# Administering a second dose of intravenous TNK in acute ischemic stroke: Rationale and design of a pilot clinical trial

**DOI:** 10.1101/2025.08.01.25332686

**Authors:** James C. Grotta

## Abstract

**Background:** A single dose of intravenous thrombolysis (IVT) is the only effective acute treatment for ischemic stroke patients who do not qualify for endovascular thrombectomy. However, at least 50% of patients treated with only IVT remain disabled. Studies suggest that incomplete recanalization and reperfusion of the distal or micro-vasculature contribute to incomplete recovery after a single dose of IVT. Single IVT doses above the approved doses produce excessive bleeding. Another strategy might be to provide a more sustained lytic effect by administering a second dose of IVT. Tenecteplase (TNK) pharmacokinetics and preliminary data from clinical trials suggest a second full dose might be given safely 45 minutes after the first dose.

**Methods:** We propose a phase 2a preliminary safety study of a second dose of TNK given 45-60 minutes after the first dose in 20 patients not responding to the first dose. Patients will be included if they qualify for and receive TNK within 3 hours of symptom onset, do not qualify for thrombectomy, sign informed consent, have NIHSS ≥ 6 45 minutes later and no bleeding on a repeat CT scan, and can receive the second TNK dose within 4.5 hours of onset.

**Results:** Primary outcome will be symptomatic intracerebral hemorrhage (type 2 parenchymal hemorrhage or parenchymal hemorrhage remote from the area of infarction with neurological deterioration) or major systemic bleeding; secondary outcomes will include other definitions of intracerebral hemorrhage and modified Rankin scale at hospital discharge and 90 days after stroke. The study will be stopped and dual full dose TNK therapy considered unsafe if 4 symptomatic or major bleeding events occur.

**Conclusion:** We propose the first study of two sequential doses of TNK in stroke patients. If successful, this study will be followed by a larger phase 2b controlled safety confirmation and pilot efficacy study.

## BACKGROUND

Intravenous thrombolysis (IVT) is an effective treatment for acute ischemic stroke (AIS) patients by dissolving the clot causing the stroke and restoring blood flow through recanalization of the affected artery. In cases where clots are located in large cerebral arteries, endovascular thrombectomy (EVT) can be employed. However, for clots located in small or medium-sized arteries, IVT remains the sole reperfusion therapy available. Patients who are eligible for “IVT-only” represent the largest cohort of AIS patients who are eligible for IVT, approximately three times larger than those who qualify for both IVT and EVT. However, in all studies of IVT for patients with AIS, at least 50% remain disabled at 90 days^1^. Although it was hoped that Tenecteplase (TNK) would prove superior to tissue plasminogen activator (tPA), results of head-to-head studies showed non-inferiority^2^. Therefore, aside from speeding IVT by improving Emergency Department metrics or deploying Mobile Stroke Units, we have not made a major advance in acute pharmacotherapy for our IVT patients since tPA was first approved in 1995. Novel approaches are needed; one such strategy might be to administer more than one dose of IVT.

Among several factors that might lead to unfavorable outcome with IVT and serve as targets for a new pharmacotherapeutic approach include failure to recanalize the artery, recanalization followed by re-occlusion, or sustained recanalization but failure to reperfuse the microcirculation^3,4^. One logical attempt to possibly prevent re-occlusion and augment reperfusion has been to combine IVT with antithrombotic or antiplatelet therapies. While there have been positive results with Argatroban and Tirofiban in Asian patients^5,6^, generally adding antiplatelets or anticoagulants to IVT has resulted in increased risk without benefit^7,8^. The recently completed Multi-arm Optimization of Stroke Thrombolysis (MOST) trial^9^ evaluated both anticoagulant and antiplatelet therapies added to IVT. Patients having National Institutes of Health Stroke Scale (NIHSS)≥6 and meeting standard criteria for IVT received either tPA or TNK within 3 hours of symptom onset. Patients were then assigned to IVT-only or IVT plus EVT cohorts based on features predicting the presence of LVO and then randomized to receive Argatroban, Eptifibatide or placebo. In both the IVT-only and IVT plus EVT cohorts, there was no benefit of either Argatroban or Eptifibatide.

A pre-specified sub-analysis of the IVT-only cohort in MOST provides clues for possible new therapeutic strategies such as repeated doses of IVT. Only 31% of patients qualifying for IVT-only harbored visible clot on computed tomography angiography (CTA) or magnetic resonance angiography (MRA), mostly in medium sized arterial branches. The remainder presumably had distal or microvascular occlusions. However, the absence of visible clots failed to predict less disabled outcome. As in previous IVT studies, 50% of patients remained disabled at 90 days with no baseline clinical or radiographic variables predictive of good outcome, including the absence of visible clot prior to treatment. The MOST trial thus re-affirmed that at least half of patients treated with IVT-only with or without visible clot and across the NIHSS spectrum will remain disabled at 90 days and suggests the importance of targeting distal occlusions and the microcirculation.

Instead of adding anti-thrombotic medications as in MOST, another strategy to target medium, distal or microcirculatory occlusions might be to provide more sustained lytic activity than can be achieved by a single dose. Supratherapeutic blood levels of lytics achieved by administering single doses above the approved 0.9 mg/kg of tPA and 0.25 mg/kg TNK have produced excessive bleeding^10,11^. Rather than higher doses, perhaps more sustained lytic activity by administering a second dose may maintain patency after successful clot lysis and also prevent clots from forming in the microcirculation. The maximal lytic effect of TNK approximately 30 minutes after bolus, and its relatively short circulatory half-life (20-25 minutes)^12^ means that a second dose might be given after the main benefit of the first dose has occurred and while brain tissue is still viable in a penumbral state which may last hours depending on collateral flow. If the first dose of TNK were given within the approved 3 hours, a second dose could still be administered after most of the first dose is dissipated and still within the 4.5-hour time window of IVT efficacy in patients whose NIHSS does not improve after the first dose.

We hypothesize that administering a second dose of TNK 45-60 minutes after the first dose might safely increase arterial recanalization/reperfusion and improve clinical outcomes compared to a single dose. We propose a study to take the first step to prove this hypothesis, e.g. to provide an initial assessment of safety.

## TRIAL DESIGN AND METHODS

The Safety of a Second Dose of TNK in Selected Acute Ischemic Stroke Patients Not Responding to the First Dose (Double Dose TNK Pilot, NCT06801054) is an open label prospective single center single arm Phase 2a study to provide a preliminary estimate of safety of two sequential doses of TNK in patients with AIS. The study was approved by the University of Texas Committee for the Protection of Human Subjects. Informed Consent will be obtained for all patients. The study is investigator initiated, designed, executed, and funded. The investigator obtained a grant from Genentech to supply the second dose of TNK. The study protocol was approved by Genentech who holds the IND. The FDA reviewed the protocol and deemed that a new IND was not necessary.

### Objectives and Outcomes

The primary objective of the study is to rule out a signal of increased bleeding. The primary outcome is symptomatic intracranial hemorrhage (sICH) defined as a type 2 parenchymal hemorrhage or a parenchymal hemorrhage remote from the area of infarction with neurological deterioration (≥4-point worsening in NIHSS)^13^, or major hemorrhage (requiring >2 units packed red blood cells), occurring within 36 hours of enrollment (e.g. at the time of the second dose of TNK). The primary outcome will be adjudicated by a senior vascular neurologist not involved in the management of the patient and experienced in the diagnosis and management of bleeding complications from IVT.

Secondary safety outcomes will include other published definitions of sICH and any bleeding. Secondary efficacy outcomes will include NIHSS and mRS at hospital discharge, and mRS at 90 days (± 14 days) after stroke onset. An exploratory efficacy objective is to compare clinical outcomes in patients receiving dual TNK therapy to propensity matched historical controls receiving only one dose of TNK extracted from our MSU database.

### Patients

Twenty patients with AIS qualifying for TNK within 3 hours of last known normal (LKN) and with disabling deficit (NIHSS ≥ 6) that persists for 45 minutes after TNK and who do not qualify for EVT will be enrolled. Inclusion and Exclusion criteria are in Table 1.

**Table 1:**
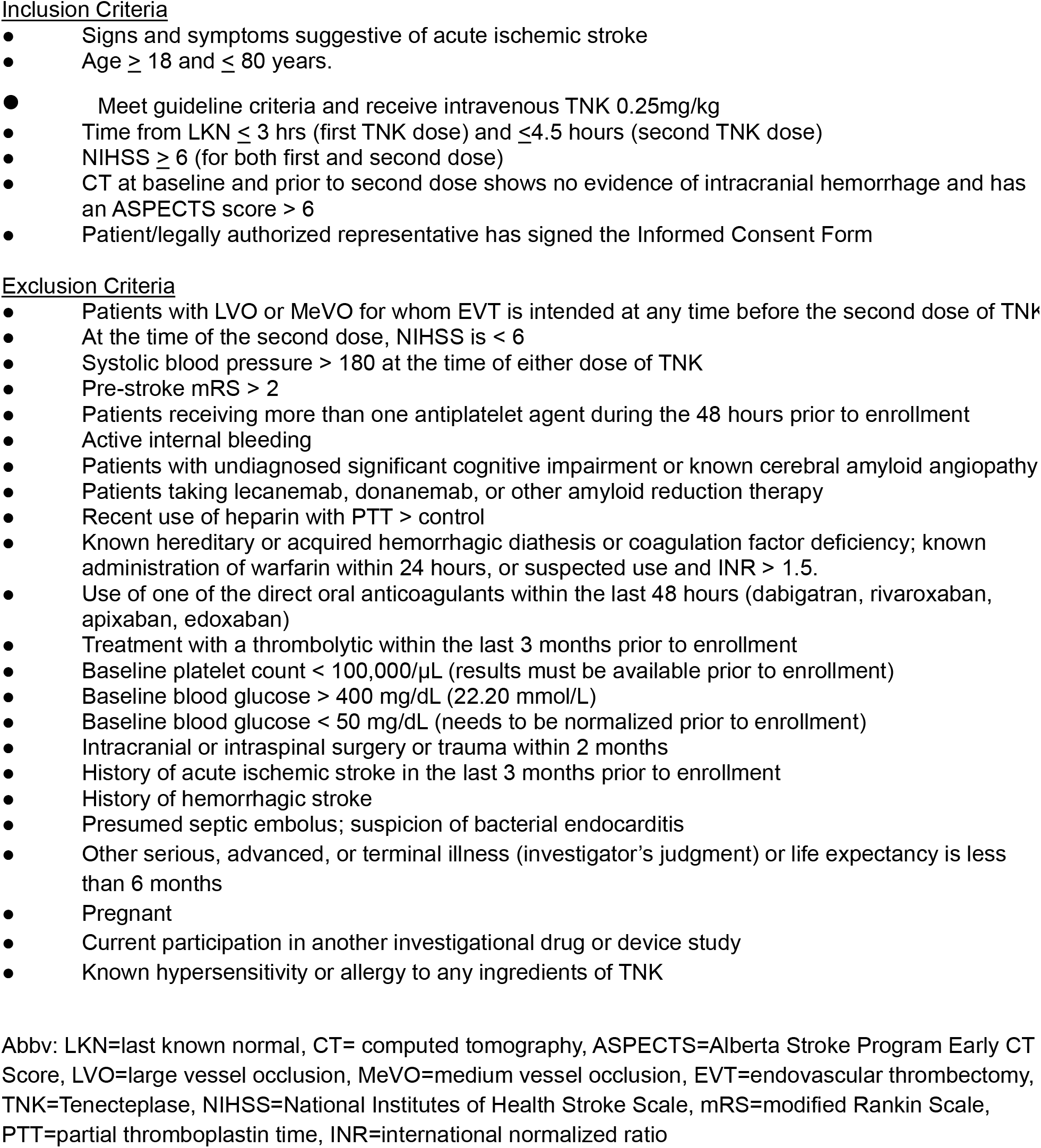
Inclusion and Exclusion criteria

Patients with medium vessel occlusions (MeVOs) can be included after consultation with the endovascular team if they are not candidates for EVT. Recent studies failed to demonstrate improved outcome with EVT in MeVO patients^14,15^ and preliminary data suggest that MeVO patients may safely respond to dual-dose IVT^16^. However, there is clinical equipoise with proximally located MeVOs, and patients with accessible clots are frequently offered EVT. Therefore, if the clot is accessible and the patient meets other criteria for EVT, they will not be included in this study.

Patients will be included based on clinical criteria rather than relying on clot identification on vascular imaging. Clinical criteria (e.g. NIHSS) have advantages to identify patients qualifying for a second dose of IVT. In MOST, 69% of patients with NIHSS ≥ 6 who qualified for IVT and not EVT did not have clot identifiable on vascular imaging and therefore probably harbored more distal occlusions^9^. Absence of visible clot did not predict increased responsiveness to IVT, and there is no reason to think that dual IVT treatment would not have benefit with more distal occlusions not seen on CTA. Furthermore, obtaining a second vascular study 1 hour after the first dose of IVT to identify persistent arterial occlusion is problematic at most clinical sites due to lack of MRA availability and excessive radiation and contrast from repeat CTA. Therefore, limiting inclusion of patients to only those with documented arterial occlusion would exclude 69% of patients, over half of whom will remain disabled after a single dose of IVT, and would severely limit study generalizability.

Our protocol will limit exposure of patients at heightened risk of bleeding or who are less likely to realize the full benefit of treatment by excluding patients with very severe strokes (LVO or CT Alberta Stroke Program Early Computed Tomography Score (ASPECTS) ≤ 6), recent use of dual antiplatelets, age > 80 years, or patients with significant cognitive impairment (to avoid patients with cerebral amyloid angiopathy). There will be no upper NIHSS cutoff since a very high NIHSS is unusual in IVT-only patients who are not candidates for EVT. In the IVT-only arm of MOST, the median baseline NIHSS was 8 (IQR 5)^9^. We will exclude patients with baseline mRS > 2 to avoid exposure of patients to uncertain risk who have no chance to achieve a non-disabled outcome.

In MOST, with similar inclusion/exclusion criteria, the mimic rate was 7% ^9^. Therefore, we expect no more than 2 mimics in our 20 enrolled patients.

### Study conduct

Patients who meet inclusion/exclusion criteria will be consented for enrollment after the first dose of TNK. After waiting 45 minutes, a non-contrast head CT scan will be obtained and read, the NIHSS repeated, and a second dose of TNK given as soon as possible thereafter (preferably within 60 minutes of the first dose) to patients who still meet inclusion/exclusion criteria provided the second dose can be given within 4.5 hours of LKN. If it is determined that the patient qualifies for EVT before the second dose is given, or if other exclusions become evident, the second dose will not be given and the patient will not be enrolled. Both TNK doses will be approximately 0.25 mg/kg, max dose 25 mg, employing FDA-approved weight-tiered dosing. The initial TNK dose may be given in a Mobile Stroke Unit or Emergency Department (ED), and the second dose in the ED. The patient will be “enrolled” only when the second TNK dose is given.

Patients will have NIHSS every 15 minutes after the first TNK dose. The NIHSS will be used to identify and exclude patients who completely respond to the first dose of IVT^17^. Early recanalization is associated with rapid clinical improvement on the NIHSS, and very low NIHSS 2 hours after dosing highly correlates with recanalization at 24 hours and eventual excellent clinical outcome^18^. Therefore, the NIHSS must be ≥ 6 at the time of the second dose. In MOST, 50% of patients with median NIHSS of 8 (IQR 5) were left disabled^9^. Therefore, if a patient improves to NIHSS < 6 between the first and second dose they will continue to be monitored in case their NIHSS worsens to ≥ 6 by 45 minutes after the first dose in which case they will be enrolled. Patients with higher NIHSS who improve after their first dose but still have NIHSS ≥ 6 45 minutes after the first dose will also be enrolled.

After enrollment and receipt of the second dose, neurological status and vital signs will be monitored every 15 minutes according to standard post-TNK routine. Patients will not be allowed antiplatelet or anticoagulant therapy for the first 24 hrs. DVT prophylaxis and single or dual antiplatelet therapy or anticoagulation may be started at 24 hrs.

Patients will receive standard AIS post-TNK management including thrombelastography (TEG) at 45 minutes and repeat CT scan at 24 ± 12 hours after the second dose of TNK or at any time there is neurologic deterioration. TEG, in particular the maximal amplitude (MA) and decrease in MA in 30 minutes (LY-30) are functional measures of circulating lytic activity and will be measured 45 minutes after the second dose when TNK levels should be waning.

## STATISTICAL CONSIDERATIONS

### Sample size

Twenty patients will be enrolled over 12 months (1.67/month). Enrollment will be stopped if 4 sICH occur, or if the independent safety monitor recommends halting the trial due to safety concerns. Using the definition to be used in this study, the rate of sICH in 617 patients receiving a single dose of IVT at our center (historical controls) is 2%^19^, and 1.8% of patients in the MOST study^9^. In the only published study to date of two sequential doses of IVT, the sICH rate was 1% in in the single IVT group and 2% in patients receiving dual doses^16^. Given the low rate of sICH with current single dose IVT, a rate > 5% with dual dosing would probably be unacceptable even if clinical outcomes were improved. If 3 sICH occur in 20 patients (15%), the 80% confidence interval is 4-26% (probably unacceptable). With 4 sICH (20%), the 80% CI is 8.6-31.4% and 90% CI is 5.4-34.6% (definitely unacceptable). Therefore, the study will be stopped if a 4th sICH occurs

### Analysis plan

Missing data: Any patient not reaching the 36-hour primary outcome follow-up point will be replaced to obtain 20 evaluable patients. Discharge mRS will be used to impute the 90 day mRS outcome in patients lost to follow up.

We will use simple proportional analysis of the confidence interval for the primary outcome, and report mean or median values for the secondary outcomes. Historical controls will be used for the exploratory clinical outcome. We will correlate TEG data with any bleeding events.

## DISCUSSION

While ours will be the first study to test two sequential doses of TNK, there are clinical trial data suggesting that administering two sequential doses of lytics is safe. In 1412 AIS pts randomized to two sequential doses of intravenous reteplase (18 mg bolus followed 30 minutes later by a second 18 mg bolus) or standard dose tPA, the dual reteplase group had no increased bleeding^20^. Intra-arterial tPA and TNK have been infused following EVT without safety concerns^21^; approximately half the patients included in one of those studies had also received standard dose IVT prior to EVT^22^. While the intra-arterial lytic dose in these trials was one-fourth to one-half of the standard IV dose, it is likely that blood levels achieved with these intra-arterial doses were higher than would occur with the same dose administered IV and might approximate full IVT dose levels.

The most relevant study to date of two sequential IVT doses evaluated full standard dose tPA followed by full standard dose TNK in patients with AIS and MeVO on baseline imaging^16^. At one hospital, 146 MeVO patients received 0.9 mg/kg IV tPA (bolus followed by 1 hr infusion) and then had repeat MRI/MRA 1 hour later. Of these, 96 had residual MeVO and were Fluid-Attenuated Inversion Recovery (FLAIR) negative and then received 0.25 mg/kg IV TNK on average 116 minutes after tPA bolus. These patients were compared by propensity matching to 148 MeVO patients receiving only IV tPA at another hospital. Patients receiving both tPA + TNK compared to patients receiving single dose tPA showed higher rates of 24 hr recanalization (82% vs 65%), and better 90 d outcome (modified Rankin Scale (mRS) 0,1 69% vs 44%). In dual-IVT patients, sICH occurred in only 2, and 1 had a psoas hematoma requiring transfusion vs 2 sICH in the tPA group. Four patients in the dual-IVT and 5 in the tPA group experienced minor bleeding. Taken together, these studies demonstrate the probable safety of dual IVT dosing and support its ability to increase recanalization in distal vessels thereby improving clinical outcome.

TNK has advantages for this dual dosing approach based on its higher fibrin specificity compared to tPA, ease of single bolus administration, and pharmacokinetics that suggest that two full doses might be safe^12^. Since the circulatory half-life of TNK is 22 min, assuming that the second TNK dose would be given approximately 60 minutes (∼2.7 half-lives) after the first dose, only ∼15% of the initial dose would remain circulating which would be equivalent to 0.038 mg/kg. Therefore, a second full dose of 0.25mg/kg would be equivalent to 0.288 mg/kg on a concentration basis which is within the FDA approved dose range.

## CONCLUSION

We describe the rationale and design for the first study of two sequential doses of TNK in patients with AIS. If successful, this study will be followed by a larger phase 2b controlled safety confirmation and pilot efficacy study.

## Data Availability

Since this is a pilot single center study, the data will not be publicaly available beyond what is published in the manuscript.

## Non-standard Abbreviations and Acronyms

IVT: intravenous thrombolysis
TNK: tenecteplase
AIS: acute ischemic stroke
EVT: endovascular thrombectomy
tPA: tissue plasminogen activator
MOST: Multi-arm Optimization of Stroke Thrombolysis
NIHSS: National Institutes of Health Stroke Scale
CTA: computed tomography angiography
MRA: magnetic resonance angiography
MeVO: medium vessel occlusion
FLAIR: fluid-attenuated inversion recovery
sICH: symptomatic intracerebral hemorrhage
mRS: modified Rankin Scale
LKN: last known normal
EVT: endovascular thrombectomy
ASPECTS: Alberta Stroke Program Early CT Score
ED: emergency department
DVT: deep venous thrombosis
TEG: thrombelastography
MA: maximum amplitude

## Disclosures

Dr Grotta receives consulting fees from Frazer, a manufacturer of Mobile Stroke Units

## Notes

Supported by grants from Genentech and the Grotta Stroke Research Foundation

### Competing Interest Statement

The authors have declared no competing interest.

### Clinical Trial

Clinical Trial identification # NCT06801054

### Funding Statement

Study drug (TNK) supplied by Genentech Funding for coordinator and pharmacy costs supplied from a local private research foundation

### Author Declarations

University of Texas Committee for the Protection of Human Subjects

## REFERENCES

1. Emberson J, Lees K, Lyden P, Blackwell L, Albers G, Bluhmki E, Brott T, Cohen G, Davis S, Donnan G et al. Effect of treatment delay, age, and stroke severity on the effects of intravenous thrombolysis with alteplase for acute ischaemic stroke: a metaanalysis of individual patient data from randomised trials. Lancet 2014; 384: 1929–35.

2. A Menon BK, Buck BH, Singh N, Deschaintre Y, Almekhlafi MA, Coutts SB, Thirunavukkarasu S, Khosravani H, Appireddy R, Moreau F, et al. AcT Trial Investigators. Intravenous tenecteplase compared with alteplase for acute ischaemic stroke in Canada (AcT): a pragmatic, multicentre, openlabel, registry-linked, randomised, controlled, non-inferiority trial. Lancet. 2022;400:161–169

3. Alexandrov AV, Grotta JC. Arterial Reocclusion in Stroke Patients Treated with Intravenous Tissue Plasminogen Activator. Neurology 2002;59:862–867.

4. Seners P, Hurford R, Tisserand M, Turc G, Legrand L, Naggara O, Mas JL, Oppenheim C, Baron JC. Is Unexplained Early Neurological Deterioration After Intravenous Thrombolysis Associated With Thrombus Extension? Stroke. 2017 48:348–352.

5. Xu J, Liu Y, Wang H, Sun R, Zhao H, Liu X, Li Y, Yang J, Zhang B, He L, et al. Effect of Argatroban Plus Dual Antiplatelet in Branch Atherosclerosis Disease: A Randomized Clinical Trial. Stroke. 2025;56:1662–1670.

6. Tao C, Liu T, Cui T, Liu J, Li Z, Ren Y, Zhao X, Xie F, Li J, Wang H, et al. ASSET-IT Investigators. Early Tirofiban Infusion after Intravenous Thrombolysis for Stroke. N Engl J Med. 2025. doi: 10.1056/NEJMoa2503678

7. van der Steen W, van de Graaf RA, Chalos V, Lingsma HF, van Doormaal PJ, Coutinho JM, Emmer BJ, de Ridder I, van Zwam W, van der Worp HB, et al. MR CLEAN-MED investigators. Safety and efficacy of aspirin, unfractionated heparin, both, or neither during endovascular stroke treatment (MR CLEAN-MED): an open-label, multicentre, randomised controlled trial. Lancet. 2022;399:1059–1069

8. Powers WJ, Rabinstein AA, Ackerson T, Adeoye OM, Bambakidis NC, Becker K, Biller J, Brown M, Demaerschalk BM, Hoh B, et al. Guidelines for the Early Management of Patients With Acute Ischemic Stroke: 2019 Update to the 2018 Guidelines for the Early Management of Acute Ischemic Stroke: A Guideline for Healthcare Professionals From the American Heart Association/American Stroke Association. Stroke. 2019;50:e344–e418. doi: 10.1161/STR.0000000000000211.

9. Adeoye O, Broderick J, Derdeyn CP, Grotta JC, Barsan W, Bentho O, Berry S, Concha M, Davis I, Demel S, et al. Adjunctive Intravenous Argatroban or Eptifibatide for Ischemic Stroke. N Engl J Med. 2024;391:810–820.

10. Brott TG, Haley EC Jr, Levy DE, Barsan W, Broderick J, Sheppard GL, Spilker J, Kongable GL, Massey S, Reed R et al. Urgent therapy for stroke. Part I. Pilot study of tissue plasminogen activator administered within 90 minutes. Stroke. 1992;23:632–40

11. Haley EC Jr, Thompson JL, Grotta JC, Lyden PD, Hemmen TG, Brown DL, Fanale C, Libman R, Kwiatkowski TG, Llinas RH, et al. TNK in Stroke Investigators. Phase IIB/III trial of TNK in acute ischemic stroke: results of a prematurely terminated randomized clinical trial. Stroke. 2010;41:707–11

12. Keyt BA, Paoni NF, Refino CJ, Berleau L, Nguyen H, Chow A, Lai J, Peña L, Pater C, Ogez J, et al. A faster-acting and more potent form of tissue plasminogen activator. Proc Natl Acad Sci U S A. 1994;91:3670–4

13. Wahlgren N, Ahmed N, Dávalos A, Ford GA, Grond M, Hacke W, Hennerici MG, Kaste M, Kuelkens S, Larrue V, et al. Thrombolysis with alteplase for acute ischaemic stroke in the Safe Implementation of Thrombolysis in Stroke-Monitoring Study (SITS-MOST): an observational study. Lancet. 2007;369:275–282.

14. Goyal M, et al ESCAPE-MeVO Investigators. Endovascular Treatment of Stroke Due to Medium-Vessel Occlusion. N Engl J Med. 2025;392:1385–1395

15. Marios-Nikos P, Alex B, Jens F, Isabel F, Jan G, Mira K, Ronen L, Paolo M, Marc R, Jeffrey L S. EnDovascular therapy plus best medical treatment (BMT) versus BMT alone for medIum distal veSsel occlusion sTroke (DISTAL): An international, multicentre, randomized-controlled, two-arm, assessor-blinded trial. Eur Stroke J. 2024;9:1083–1092

16. Chausson N, Olindo S, Laborne FX, Aghasaryan M, Renou P, Soumah D, Debruxelles S, Altarcha T, Poli M, L’Hermitte Y, et al. Second-dose intravenous thrombolysis with TNK in alteplase-resistant medium-vessel-occlusion strokes: A retrospective and comparative study. Eur Stroke J. 2024;9:943–951.

17. Mikulik R, Ribo M, Hill MD, Grotta JC, Malkoff M, Molina C, Rubiera M, Delgado-Mederos R, Alvarez-Sabin J, Alexandrov AV; CLOTBUST Investigators. Accuracy of serial National Institutes of Health Stroke Scale scores to identify artery status in acute ischemic stroke. Circulation. 2007;115:2660–5.

18. Kharitonova T, Mikulik R, Roine RO, Soinne L, Ahmed N, Wahlgren N; Safe Implementation of Thrombolysis in Stroke Investigators. Association of early National Institutes of Health Stroke Scale improvement with vessel recanalization and functional outcome after intravenous thrombolysis in ischemic stroke. Stroke. 2011;42:1638–43.

19. Grotta JC, Yamal JM, Parker SA, Rajan SS, Gonzales NR, Jones WJ, Alexandrov AW, Navi BB, Nour M, Spokoyny I, et al. Prospective, Multicenter, Controlled Trial of Mobile Stroke Units. N Engl J Med. 2021;385:971–981. Erratum in: N Engl J Med. 2023;388:2495-2496.

20. Li S, Gu HQ, Li H, Wang X, Jin A, Guo S, Lu G, Che F, Wang W, Wei Y, et al. Reteplase versus Alteplase for Acute Ischemic Stroke. N Engl J Med. 2024;390:2264–2273

21. Jiang X, Zhao Z, Zhang Y, Luo W, Zheng K, Zhang M, Li E, Lang H, Wang J, Zhou C, He L. Intra-Arterial Thrombolysis Following Endovascular Recanalization for Large Vessel Occlusion Stroke: A Systematic Review and Meta-Analysis. Neurology. 2025;105:e213842. doi: 10.1212/WNL.0000000000213842

22. Renú A, Millán M, San Román L, Blasco J, Martí-Fàbregas J, Terceño M, Amaro S, Serena J, Urra X, Laredo C, et al. Effect of Intra-arterial Alteplase vs Placebo Following Successful Thrombectomy on Functional Outcomes in Patients With Large Vessel Occlusion Acute Ischemic Stroke: The CHOICE Randomized Clinical Trial. JAMA. 2022;327:826–835

